# Longitudinal changes of iron in the substantia nigra of early-stage Parkinson’s disease

**DOI:** 10.64898/2026.06.02.26354736

**Authors:** Guangwei Du, Christopher Sica, Sol De Jesus, Lan Kong, Richard B. Mailman, Xuemei Huang

**Affiliations:** Department of Neurology, Penn State Milton S. Hershey Medical Center, Hershey PA 17033; Department of Radiology, Rush University, Chicago, IL 60612; School of Public Health Sciences, Pennsylvania State College of Medicine, Hershey, PA 17033; Department of Pharmacology, University of Virginia, Charlottesville, VA 22908; Department of Neurology, University of Virginia, Charlottesville, VA 22908

**Author notes:** ***Corresponding Author:*** Guangwei Du, MD, PhD, Department of Neurology, Penn State Milton S. Hershey Medical Center, 500 University Drive H037, Hershey PA, 17033.

**Keywords:** longitudinal, substantia nigra, brain iron, susceptibility MRI, Parkinson’s disease

## Abstract

**Background:** Higher iron occurs in the substantia nigra in Parkinson’s disease (PD), but its behavior over time is poorly understood.

**Objective:** To determine if nigral iron measured by quantitative susceptibility MRI can track progression of early stages of PD.

**Methods:** Thirty-four early-stage PD patients (disease duration < 4 yrs) and 35 matched controls were recruited. Iron content in the substantia nigra and related regions was obtained using quantitative susceptibility mapping with zero-referencing at both baseline and one-year follow-up, along with clinical metrics.

**Results:** Of five clinic metrics and five regional iron measures, only MDS-UPDRS II (p = 0.046) and SNc iron (p = 0.004) showed significant changes for PD subjects in the one-year epoch. The changes of SNc iron also were correlated with changes of MDS-UPDRS II (p = 0.005).

**Conclusion:** Nigral iron content measured by zero-referencing is sensitive for tracking disease progression in early-stage PD.

## Introduction

Parkinson’s disease (PD) is marked pathologically by dopamine neuron loss in the substantia nigra (SN) pars compacta (SNc) of the basal ganglia^1,2^ and by increased iron in the SN.^3-11^ The causes and effects of the higher nigral iron in PD are not understood. Because excessive iron has been linked to higher oxidative stress (and ultimately dopamine neuron death in the SN),^12-16^ higher iron content in PD has been hypothesized to play a role in PD etiology.^17-19^ Few studies, however, have focused on longitudinal changes and whether accumulation of iron in the SNc is a progression marker for PD, particularly in early stages.

Emerging MRI technology has enabled *in vivo* quantification of brain iron content in humans.^11,20,21^ The data from use of susceptibility MRI correlates well with post-mortem chemical measurement of iron in deep gray matter regions.^22,23^ Studies from our group and others have used both apparent transverse relaxation rate (R2*) and newer quantitative susceptibility mapping (QSM) methods to quantify iron in the SN of PD patients.^8,20,24-26^ The QSM technique used previously studies, however, had a significant limitation. There is an arbitrary constant shift in the obtained QSM values, because the measured tissue susceptibility is invariant with any uniform shift in the true underlying susceptibility distribution..^27^ This can be extremely problematic when using QSM longitudinally. For correction, a zero-referencing technique can subtract a mean QSM value from the lateral ventricle area where the cerebrospinal fluid is the main source of signal.^27^ This technique also reduces artifacts near the ventricles.

Thus, we studied early-stage PD patients using zero-referencing quantitative susceptibility MRI methods to investigate longitudinal changes in nigral iron and other related regions. We tested the following hypotheses: 1) nigral iron measured by QSM can track progression in early-stage PD; and 2) the detected nigral iron change is associated with meaningful clinical progression.

## Methods

### Study design and subjects

Thirty-four PD patients were recruited from a tertiary movement disorders clinic, and 35 controls were recruited from spouses and the local community. PD diagnosis was confirmed based on the Movement Disorder Society criteria.^28^ To capture early -stage changes, only PD patients with < 4 years of disease duration were recruited. Disease duration for PD patients was defined based on the time since first documented PD diagnosis. All participants were free of major medical issues or neurological conditions other than PD. Demographic and clinical characteristics including the Movement Disorder Society Unified PD Rating Scale parts I, II, and III (MDS-UPDRS-I, -II, -III); the Geriatric Depression Scale (GDS); and the Montreal Cognitive Assessment (MoCA) were obtained from all participants while patients were on optimized antiparkinsonian medications (ON state). All clinical metrics and MRI were obtained at both baseline and one-year follow-up. Detailed demographic data are provided in Table 1. All participants gave written informed consent that was in accordance with the Declaration of Helsinki, and the protocol was approved by the Penn State Hershey Institutional Review Board.

**Table 1.**
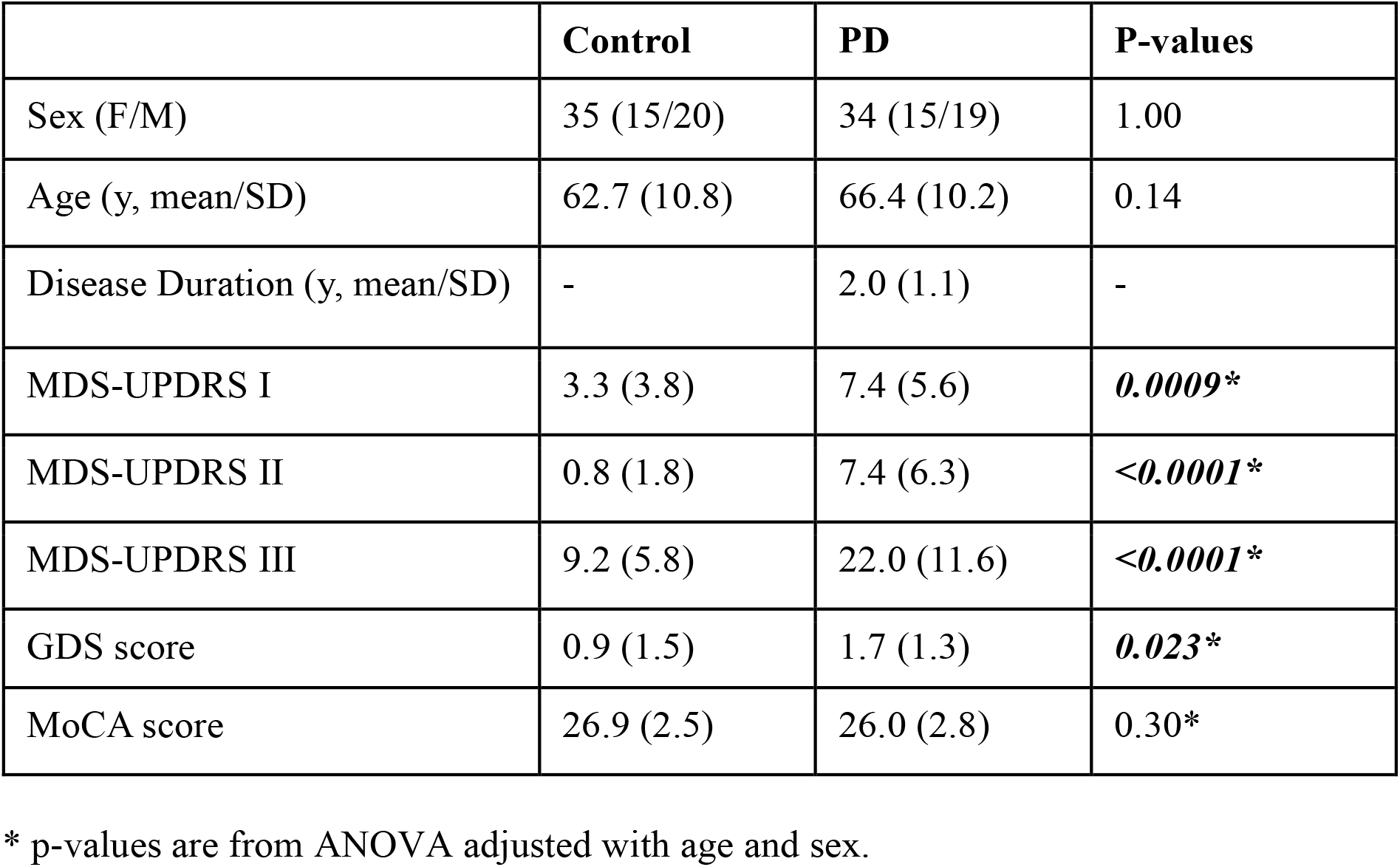
Demographics.

### MRI Image acquisition and analysis

All images were acquired on a 3T Siemens PrismaFit scanner (Siemens Healthineers, Erlangen, Germany) with a 20-channel coil. An eight-echo multi-gradient-echo sequence was used for QSM reconstruction. Quantitative susceptibility maps (QSM) were generated using morphology enabled dipole inversion using an automatic uniform cerebrospinal fluid zero reference (MEDI+0) with a nonlinear formation of the magnetic field to source.^27^

### Regions-of-interest segmentation

Regions-of-interest (ROIs) included in the study were basal ganglia-related structures: substantia nigra pars compacta (SNc), putamen (PUT), caudate (CN), globus pallidus (GP), and red nucleus (RN).^29-32^ The ROIs were segmented using semi-automatic atlas-based parcellation to improve the accuracy and repeatability of the segmentation.^11^ Finally, mean QSM values from each ROI were calculated for individual subjects. [Detailed acquisition parameters and image analysis method are in Supplemental Material]

### Statistical analysis

Demographic data were compared between PD patients and controls using the Fisher’s exact test for sex and the two-tailed Student’s t-test for age. MDS-UPDRS-I, -II, -III, MoCA, GDS, and UPSIT were compared between patients and controls using one-way analyses-of-covariance (ANCOVA) with adjustments of age and sex at baseline. Longitudinal changes of clinical metrics and regional QSM values were assessed using linear mixed effect modeling with Visit, Group, and Visit × Group as main effects off interests, and age, sex as covariates. Bonferroni method was used for multiple comparison correction for the primary outcomes (i.e., for five Visit × Group effects of clinical metrics or 5 regional QSM values is considered at a time). The p-value < 0.01 (0.05 ÷ 5) was set as the threshold for significance after correction. The correlations between clinical and MRI measures were evaluated using partial Pearson coefficients with age and sex as covariates. All statistical analyses were performed using SAS 9.4 (SAS Institute Inc., Cary, NC).

## Results

### Demographic and clinical information

Demographic and clinical data for each group are summarized in Table 1. There was no significant difference between PD patients and controls in terms of age or sex. As expected, at baseline PD patients showed worse clinical measures in all metrics except the MoCA score.

### Longitudinal analysis of clinical metrics and regional QSM values using linear mixed-effect models

For clinical metrics, only MDS-UPDRS II showed a significant longitudinal effect indicated by Visit × Group effect (p = 0.046). It, however, was not significant after multiple comparison correction. Among five regional QSM values, only SNc showed significant Visit, Group, and Visit × Group effects. Both Group and Visit × Group effects survived after multiple comparison correction. MDS-UPDRS II showed significant differences at both baseline (p < 0.0001) and one-year follow-up (p < 0.0001); the SNc QSM values, however, showed only significant differences at one-year follow-up (p = 0.001) but not at baseline (p = 0.207). The longitudinal changes patterns were shown in Figure 1A and 1B. The post-hoc power analyses showed that the MDS-UPDRS II has an effect size = 0.42 for the longitudinal changes over a one-year period. This would require 154 patients to achieve 0.95 power if used in a clinical trial to detect longitudinal changes. Conversely, the SNc QSM value has an effect size = 1.14 for the longitudinal changes over one-year period. It would require only 26 patients to achieve 0.95 power when detecting longitudinal changes of PD patients.

**Figure 1.**
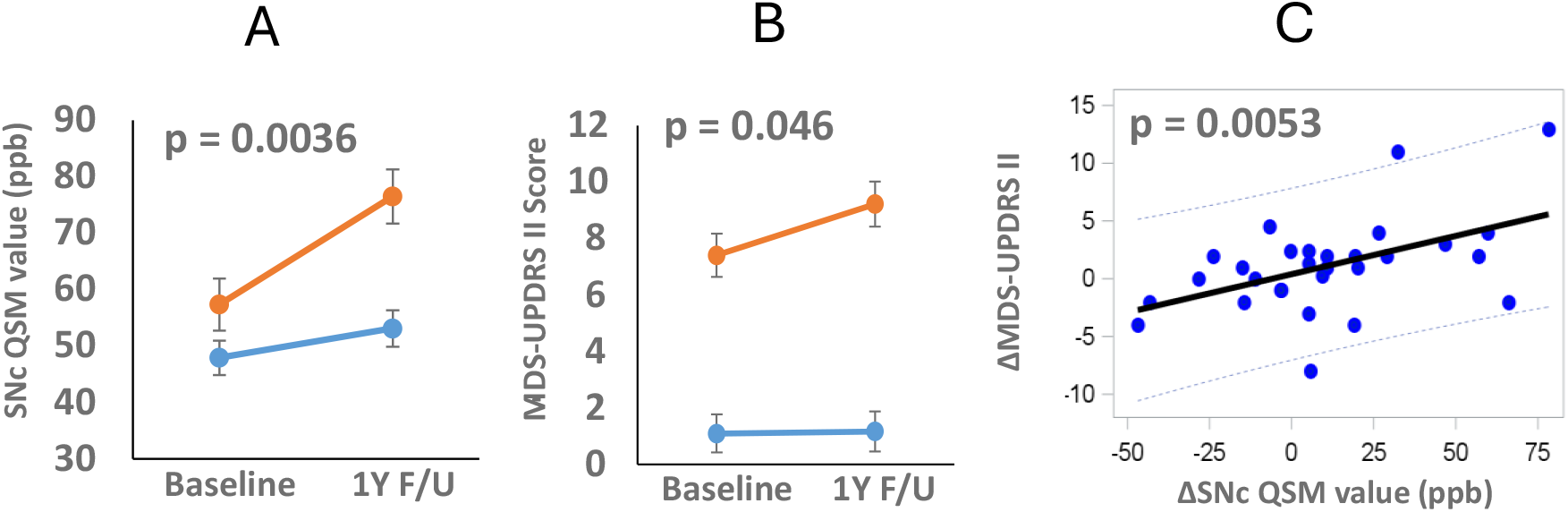
Comparison of longitudinal progression of SNc QSM values (A) and MDS-UPDRS II (B) between PD (orange) and controls (light blue). The mean and standard error bars are from linear mixed model after controlling for baseline age and sex. The p-values are the Group × Visit effects from linear mixed-effects models. C is scatterplot showing the correlation between 1 year change of SNc QSM and MDS-UPDRS II scores. The p-values are from partial Pearson correlation coefficient with baseline age and sex as covariates.

### Correlation between clinical metrics and regional QSM values

At baseline, the CN QSM values were correlated with MDS-UPDRS I, II, and III, the GP QSM values are correlated with MoCA. Longitudinally, the 1-year change of SNc QSM values is correlated with the 1-year changes of MDS-UPDRS I (r = 0.462, p = 0.023) and II scores (r = 0.536, p = 0.0053). The change in PUT QSM values is correlated with the change in GDS scores (r = 0.566, p = 0.003). (See supplemental Table 2 and 3). The significant correlation between MDS-UPDRS II changes and SNc QSM emphasizes the clinical relevance of SNc QSM values as a novel and sensitive progression marker for PD. (See Figure 1C)

## Discussion

We examined whether regional brain iron content can track disease progression in early-stage PD patients using a novel zero-referencing QSM technique, along with conventional clinical metrics. Our study is the first to demonstrate that nigral iron measured by zero-referencing QSM can track disease-specific progression in early-stage PD over 1-year epoch. We also found that MDS-UPDRS II, a self-reported motor symptom severity metric, also can be used to track disease progression in the early-stage of PD, but with a much smaller effect size. These findings were corroborated with a significant correlation between SNc QSM changes and MDS-UPDRS II changes over a one-year period.

### Nigral iron content and MDS-UPDRS II track PD progression at early-stage

Few studies have focused on discovering progression markers for early stages of PD. Previously, using a larger cohort, we reported that both R2* and QSM can detect mid- to late-stage nigral iron changes over an 18-month epoch.^11^ Recently, Gaurav et al reported that posteroventral SN QSM values in PD patients progressed during a 4-year epoch.^33^ Free-water diffusion also has been demonstrated to detect longitudinal changes of substantia nigra of PD patients at early-stage but with a smaller effect size, thus requiring a larger sample size (176 at 50% effect of the tested drug)^34^ for use in therapeutic trials. Newer QSM zero-referencing technique is an elegant and simple improvement that eliminates the arbitrary constant shifting of QSM values, which is due to the dipole kernel used for field-to-source inversion having a singularity at k-space center.^27^ It allows QSM to detect longitudinal changes in repeated measures of the same patient with much higher accuracy.^27^ Using this technique, we have demonstrated that the SNc QSM can be a sensitive progression marker for early-stage PD, with a large effect size, thus a much smaller, sample size can be used in future therapeutic trials.

As noted above, higher SNc iron has been suggested to play a role in PD etiology.^12-19^ Previous work from our group with a larger cohort and a drug naïve group showed that nigral iron does not increase until 2-3 years after diagnosis and drug use.^10^ Our data showed no significant difference in iron content between PD and control at baseline, a result consistent with a recent report from another group.^35^ One plausible explanation is that the iron increases are a result of the medication(s) that is/are the standard of care for PD and that start after diagnosis.^10,36,37^ Further research is needed to resolve this conundrum by using a large cohort study of de novo PD patients.

### Limitations

Our study has a few limitations. First, the sample size is relatively small compared to recent neuroimaging studies using susceptibility MRI.^33^ This is, however, offset by the larger effect size we have found. Second, due to the nature of MRI, the QSM signal merely measures bulk susceptibility effects within a voxel. Despite previous work having suggested that the iron bound to ferritin might dominate the QSM signal, other substances, such as deoxygenated blood, transferrin, myelin, and calcium also contribute to bulk susceptibility within the same voxel.^22^ The neurobiological underpinning of nigral QSM changes is, therefore, complex.

In summary, the SNc iron measured by zero-referencing QSM provides a more sensitive imaging biomarker for disease modification assessments targeting iron or related PD pathologies. Validation of this finding and comparison with other potential progression markers with a large cohort is warranted.

## Data Availability

All data produced in the present study are available upon reasonable request to the authors.

## Acknowledgements

We express gratitude to all the participants who volunteered for this study and study personnel who contributed to its success. We also thank our diligent research coordinators Autumn Collier and Hong Wang who were key to the success of this project.

## Author contributions

1. Research project: A. Conception, B. Organization, C. Execution;

2. Statistical Analysis: A. Design, B. Execution, C. Review and Critique;

3. Manuscript: A. Writing of the first draft, B. Review and Critique;

Guangwei Du: 1A, 1B, 1C, 2A, 2B, 3A, 3B.

Christopher Sica: 1C, 3B.

Sol De Jesus: 1A, 1B, 1C, 3B.

Lan Kong: 2A, 2C, 3B.

Richard Mailman: 1A, 2C, 3B.

Xuemei Huang: 1A, 1B, 1C, 3B.

