## Supplemental Data for "Longitudinal changes of iron in the substantia nigra of early-stage Parkinson’s disease"

**MRI acquisition and ROI segmentation.**

For each participant, T1-weighted, T2-weighted, and multi-gradient-echo MR images were acquired on a 3T Siemens PrismaFit scanner (Siemens Healthineers, Erlangen, Germany) with a 20-channel coil. The T1- and T2-weighted images are used as anatomical references. A magnetization-prepared rapid acquisition gradient echo sequence was used to obtain T1-weighted images with repetition time/echo time = 1540/2.34 milliseconds; field of view = 256×256; slice thickness, 1 mm (with no gap); and slice number = 176. A 3-dimensional T2-weighted SPACE (sampling perfection with application optimized contrast using different angle evolution) sequence was used to obtain T2-weighted images with repetition time/echo time = 2500/316 milliseconds and the same spatial resolution settings as the T1-weighted images. T2*-weighted images were acquired using a multi-gradient-echo sequence with 8 echoes (echo times ranging from 6.2 to 49.6 milliseconds); and repetition time = 55 milliseconds; flip angle = 15°; field of view = 240×240; matrix = 256×256; slice thickness = 2 mm; slice number = 64; and voxel size = 0.9×0.9×2 $mm^3$. All images were inspected offline and deemed free of severe artifacts or any major structural abnormalities.

An atlas-based semiautomatic segmentation approach was used to segmented ROIs in the study. Namely, In step 1, T1- and T2-weighted images from all participants were used to construct a cohort-specific template using an unbiased atlas construction algorithm in the Advanced Normalization Tools (ANTS) package.[1,2] Since most of these structures have high iron content and are seen best in T2-weighted images, the ROIs were defined manually on the constructed T2-weighted template according to previous studies.[3,4] Supplemental Figure 1 shows the exact location of these five structures. In step 2, an atlas-based segmentation pipeline (AutoSeg v3.0; University of North Carolina Neuro Image Analysis Laboratory) generated the segmentation in individual space based on the T2-weighted images. In step 3, automatic segmentation results from all subjects were inspected visually and corrected manually by a rater blinded to group information. In step 4, an affine registration algorithm was used to bring the ROIs from T2-weighted images to the QSM images. Finally, mean QSM values from each ROI (including both left and right sides) were calculated for individual subjects.

**Supplemental Figure 1. Definition of regions of interests (ROI) on T2w template image.**

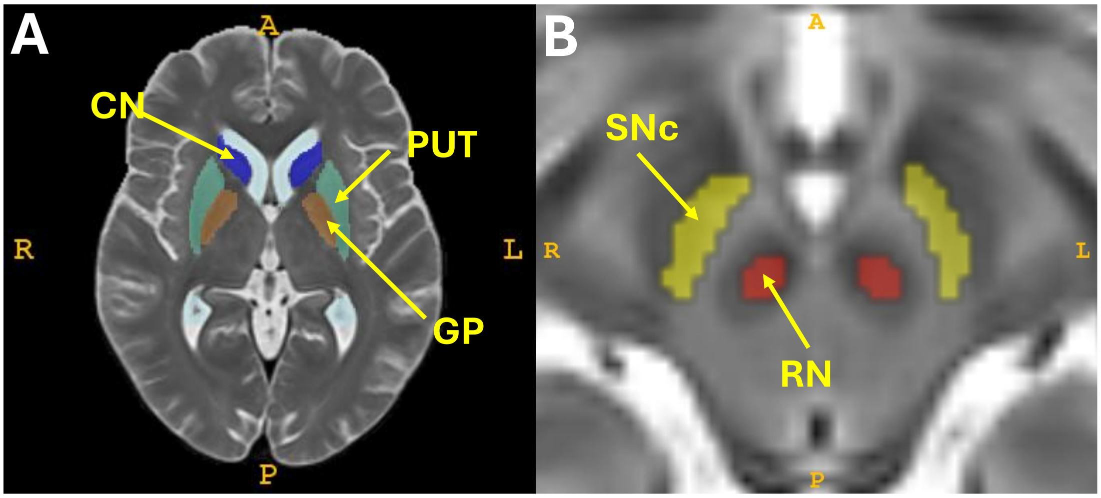

**Supplemental Table 1. Longitudinal analysis results with linear mixed models.**

| | Visit | Group | Visit × Group | Age | Sex |
|---|---|---|---|---|---|
| Clinical metrics: | | | | | |
| MDS-UPDRS I | 0.894 | **0.0007*** | 0.567 | 0.675 | 0.926 |
| MDS-UPDRS II | **0.032** | **<0.0001*** | **0.046** | 0.172 | 0.187 |
| MDS-UPDRS III | 0.270 | **<0.0001*** | 0.328 | **0.011** | 0.365 |
| GDS scores | 0.625 | **0.0063** | 0.916 | 0.805 | 0.259 |
| MoCA scores | 0.767 | **0.018** | 0.079 | **0.026** | 0.916 |
| QSM measures: | | | | | |
| SNc | **0.0028*** | **0.0008*** | **0.0036*** | 0.741 | 0.259 |
| RN | 0.470 | 0.092 | 0.402 | 0.206 | 0.737 |
| PUT | 0.626 | 0.159 | 0.619 | **0.013** | 0.822 |
| CN | 0.840 | 0.182 | 0.760 | 0.470 | 0.420 |
| GP | 0.916 | 0.114 | 0.271 | 0.989 | 0.108 |

The p-values are from mixed effects models. * indicates significance after Bonferroni correction.

**Supplemental Table 2. Correlation between regional QSM values and clinical metrics at baseline.**

| | | SNc | RN | PUT | CN | GP |
|---|---|---|---|---|---|---|
| **MDS-UPDRS I** | r | 0.268 | 0.105 | 0.234 | ***0.447*** | 0.124 |
| | p | 0.176 | 0.602 | 0.241 | ***0.019*** | 0.538 |
| **MDS-UPDRS II** | r | -0.072 | -0.067 | 0.302 | ***0.423*** | -0.149 |
| | p | 0.722 | 0.741 | 0.126 | ***0.028*** | 0.458 |
| **MDS-UPDRS III** | r | 0.139 | 0.093 | -0.101 | ***0.408*** | 0.247 |
| | p | 0.489 | 0.646 | 0.615 | ***0.034*** | 0.213 |
| **GDS score** | r | 0.186 | 0.043 | 0.176 | 0.130 | 0.125 |
| | p | 0.352 | 0.830 | 0.380 | 0.519 | 0.536 |
| **MoCA score** | r | -0.140 | -0.060 | -0.199 | -0.189 | ***-0.392*** |
| | p | 0.486 | 0.764 | 0.320 | 0.344 | ***0.043*** |

**Supplemental Table 3. Correlation between 1-year changes of regional QSM values and clinical metrics.**

| | | SNc | RN | PUT | CN | GP |
|---|---|---|---|---|---|---|
| **MDS-UPDRS I** | r | ***0.462*** | 0.256 | 0.229 | 0.303 | 0.233 |
| | p | ***0.023*** | 0.227 | 0.282 | 0.150 | 0.273 |
| **MDS-UPDRS II** | r | ***0.536*** | 0.290 | 0.193 | 0.154 | 0.042 |
| | p | ***0.005**** | 0.159 | 0.356 | 0.463 | 0.842 |
| **MDS-UPDRS III** | r | 0.242 | 0.047 | -0.238 | -0.188 | -0.061 |
| | p | 0.244 | 0.822 | 0.251 | 0.368 | 0.773 |
| **GDS score** | r | -0.024 | -0.134 | ***0.566*** | 0.351 | -0.255 |
| | p | 0.911 | 0.523 | ***0.003**** | 0.085 | 0.219 |
| **MoCA score** | r | 0.141 | 0.232 | -0.029 | 0.003 | -0.011 |
| | p | 0.501 | 0.266 | 0.891 | 0.989 | 0.958 |

The p-values are from partial Pearson correlation analyses. * indicates significance after Bonferroni correction.
